# Nonlinear biomarker interactions in conversion from Mild Cognitive Impairment to Alzheimer’s disease

**DOI:** 10.1101/19002378

**Authors:** Sebastian G. Popescu, Alex Whittington, Roger N. Gunn, Paul M. Matthews, Ben Glocker, David J Sharp, James H Cole, for the Alzheimer’s Disease Neuroimaging Initiative

## Abstract

The multi-faceted nature of Alzheimer’s disease means that multiple biomarkers (e.g., amyloid-β, tau, brain atrophy) can contribute to the prediction of clinical outcomes. Machine learning methods are a powerful way to identify the best approach to this prediction. However, it has been difficult previously to model nonlinear interactions between biomarkers in the context of predictive models. This is important as the mechanisms relating these biomarkers to the disease are inter-related and nonlinear interactions occur. Here, we used Gaussian Processes to model nonlinear interactions when combining biomarkers to predict Alzheimer’s disease conversion in 48 mild cognitive impairment participants who progressed to Alzheimer’s disease and 158 stable (over three years) people with mild cognitive impairment. Measures included: demographics, APOE4 genotype, CSF (amyloid-β42, total tau, phosphorylated tau), neuroimaging markers of amyloid-β deposition ([18^F^]florbetapir) or neurodegeneration (hippocampal volume, brain-age). We examined: (i) the independent value each biomarker has in predicting conversion; and (ii) whether modelling nonlinear interactions between biomarkers improved prediction performance.

Despite relatively high correlations between different biomarkers, our results showed that each measured added complementary information when predicting conversion to Alzheimer’s disease. A linear model predicting MCI group (stable versus progressive) explained over half the variance (R^2^ = 0.51, *P* < 0.001); the strongest independently-contributing biomarker was hippocampal volume (R^2^ = 0.13). Next, we compared the sensitivity of different models to progressive MCI: independent biomarker models, additive models (with no interaction terms), nonlinear interaction models. We observed a significant improvement (*P* < 0.001) for various two-way interaction models, with the best performing model including an interaction between amyloid-β-PET and P-tau, while accounting for hippocampal volume (sensitivity = 0.77).

Our results showed that closely-related biomarkers still contribute uniquely to the prediction of conversion, supporting the continued use of comprehensive biological assessments. A number of interactions between biomarkers were implicated in the prediction of Alzheimer’s disease conversion. For example, the interaction between hippocampal atrophy and amyloid-deposition influences progression to Alzheimer’s disease over and above their independent contributions. Importantly, nonlinear interaction modelling shows that although for some patients adding additional biomarkers may add little value (i.e., when hippocampal volume is high), but for others (i.e., with low hippocampal volume) further invasive and expensive testing is warranted. Our Gaussian Processes framework enables visual examination of these nonlinear interactions, allowing projection of individual patients into biomarker ‘space’, providing a way to make personalised healthcare decisions or stratify subsets of patients for recruitment into trials of neuroprotective interventions.

## Introduction

Rapid increases in the prevalence of Alzheimer’s disease over the 21st century are predicted (Brookmeyer *et al*., 2007), hence there is a pressing need to develop disease modifying treatments. Research diagnostic criteria have been proposed, reflecting key biological facets of Alzheimer’s such as cerebral amyloid-β deposition, altered levels of proteins in CSF and neurodegeneration (Sperling *et al*., 2011; Dubois *et al*., 2014). It is hoped that these biological characteristics can be measured in people prior to symptoms manifesting, identifying at-risk individuals and enabling interventions to be targeted at slowing disease progression and delaying symptom onset.

This goal is complicated by the highly heterogeneous nature of the Alzheimer’s population (Tatsuoka *et al*., 2013). For example, although the accumulation of amyloid-β plaques is a defining pathological feature of Alzheimer’s, many cognitively-normal older adults also have elevated amyloid-β plaque levels (Villemagne *et al*., 2011). Seemingly, amyloid-β deposition is not solely sufficient to cause dementia (Aizenstein *et al*., 2008). Furthermore, while CSF tau was found to positively correlate with severity of cognitive impairment (Shaw *et al*., 2009), increased CSF tau appears to indicate neuronal injury and neurodegeneration in different diseases (Schoonenboom *et al*., 2011). Indeed, a broad range of potential biomarkers are available for Alzheimer’s. This includes hippocampal volume, whole-brain volume and as well as apparent ‘brain-age. Previous work has shown that that having an older-appearing brain is associated with conversion to Alzheimer’s within three years, and is more statistically informative than hippocampal volume (Franke *et al*., 2010; Franke and Gaser, 2012; Gaser *et al*., 2013). We have previously used the brain-age paradigm to demonstrate abnormal brain-ageing after a traumatic brain injury, in treatment resistant epilepsy and Down’s syndrome (Cole *et al*., 2015; Cole *et al*., 2017a; Pardoe *et al*., 2017). Brain-age also relates to cognitive performance and mortality risk in older adults from the general population (Cole *et al*., 2018), suggesting that this metric reflects something of the brain’s sensitivity to more general health. The range of available biomarkers indicates the biological heterogeneity of Alzheimer’s and more work is needed to incorporate this heterogeneity into predictive models of disease progression. This should improve specificity and better enable treatment decisions to be made at the individual level, to aid in clinical practice and evaluate potential treatments.

To improve specificity in the use of biomarkers for staging Alzheimer’s, multiple measures are likely to be necessary. This way, complementary information from different biological sources can be combined to build a more comprehensive picture of the underlying diseases processes, and capture disease heterogeneity more accurately. An essential consideration when combining these multiple sources of information is how they interact. For instance, while amyloid-β deposition may precede and potentially drive subsequent neurodegeneration in some instances, the magnitude of amyloid-β deposition is likely to be key. An individual may be ‘positive’ for amyloid-β on a PET scan but remain below some latent threshold for neuronal or glial loss. Once over that threshold, neurodegeneration might occur, though if they remain below this threshold, neurodegeneration may be driven by other factors, or not occur at all. This accords with Jack and colleagues (2016) proposed multi-state transition model, with two distinct pathways to Alzheimer’s; one where cerebral amyloid-β deposition occurs prior to neurodegeneration, and one where neurodegeneration occurs prior to amyloid-β deposition. In this second pathway, individuals may never accumulate sufficient amyloid-β to cross the threshold to amyloid-mediated neurodegeneration. However, they may have been exposed to negative genetic or environmental factors, not necessarily Alzheimer’s related, that have influenced the rate of age-associated change in brain structure. On this back-drop of poorer brain health, potentially only minimal Alzheimer’s-specific pathology is necessary to drive disease progression. The presence of such thresholds in how two biological processes interact is inherently nonlinear, however, nonlinear relationships are under-studied in neurology and remain in the domain of systems biology, for example in modelling cell signalling networks (Sung and Hager, 2012). Nonlinear models that incorporate thresholds, plateaus and other complex patterns are theoretically better positioned to detect such relationships, and machine learning analysis offers a range of tools for modelling nonlinearities.

Machine learning is an increasingly popular approach to predict conversion from MCI to Alzheimer’s using biomarkers (Sarica *et al*., 2017; Pellegrini *et al*., 2018). Machine learning places emphasis on out-of-sample prediction and is readily able to incorporate high-dimensional data, hence is well-suited to making individualised predictions of disease outcomes. However, only a limited number of studies have specifically modelled interactions between biomarkers and crucially, these have only been linear interactions (Fortea *et al*., 2014; Li *et al*., 2014; Caroli *et al*., 2015; Pascoal *et al*., 2017a; Pascoal *et al*., 2017b; Bilgel *et al*., 2018). For example, Pascoal and colleagues (2017b) showed that being positive for both amyloid-β-PET and CSF p-tau was associated greater cognitive decline and greater risk of disease progression in people with MCI, then being positive for either biomarker in isolation. However, when dichotomising the continuous measures of amyloid-β-PET and CSF p-tau into positive or negative, it is not possible to detect more complex relationships between biomarkers. Hence, this motivates research into alternative methods that enable both linear and nonlinear interactions to be modelled.

One approach to modelling nonlinear interactions is Gaussian Processes, a family of models that assigns a nonparametric Gaussian smoothing function to each biomarker (Figure 1). These ‘kernels’ can then be combined, allowing effects of non-linearities in both additive (i.e., main effects) and interactions to be modelled. Here, we tested the hypothesis that the conversion from MCI to Alzheimer’s can be better predicted by modelling biomarkers interactions (linear or non-linear), than by using the additive sum of independent effects on disease progression.

**Figure 1.**
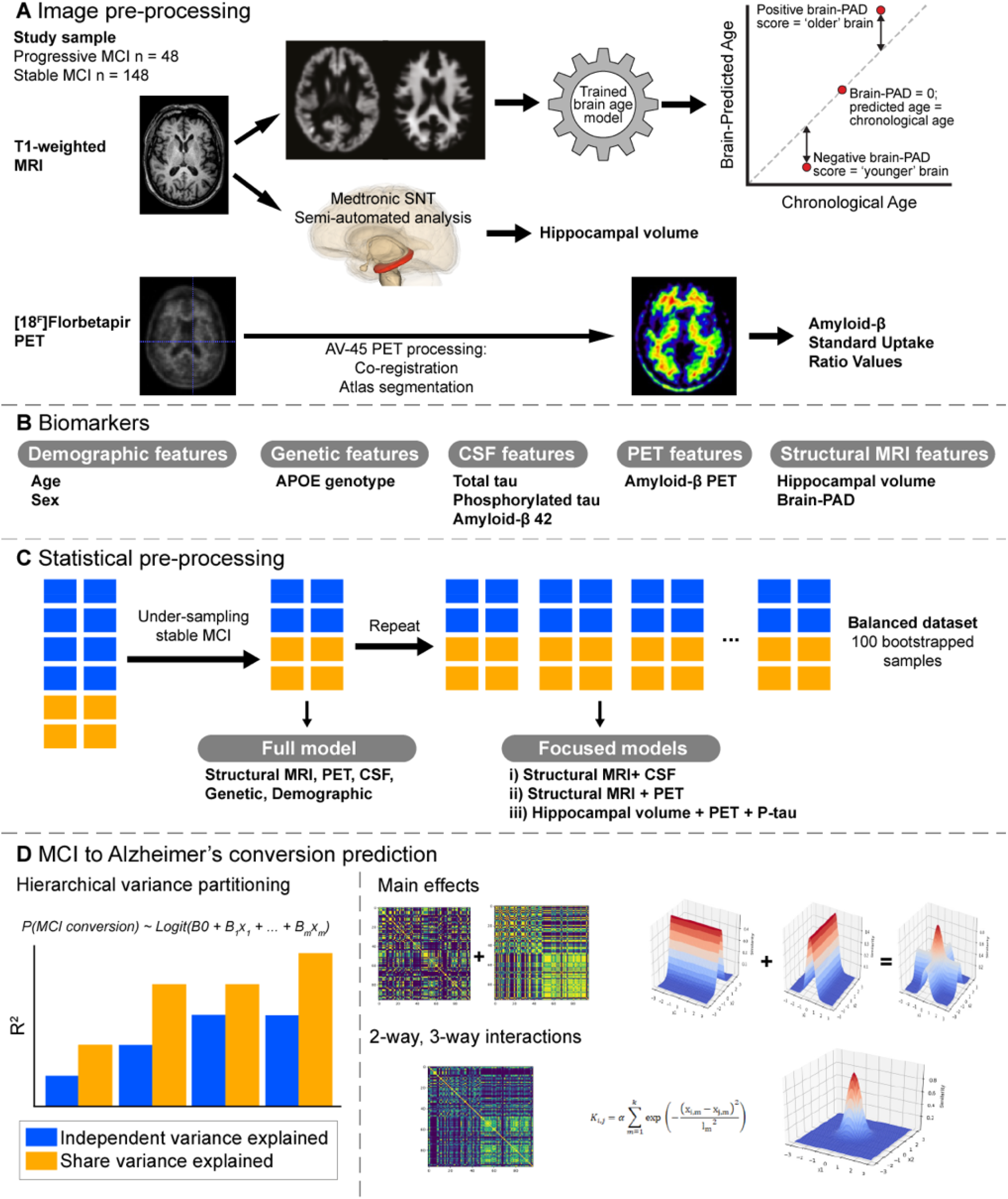
Overview of study methods. A) Raw T1-weighted MRI scans are pre-processed via the DARTEL pipeline in SPM12 to obtain grey matter and white matter volume maps. These are fed into a pretrained Gaussian Processes Regression based “brain-age” prediction software to arrive at an estimation of Brain-PAD, which is the difference between chronological age and neuroimaging-predicted “brain-age”. B) CSF features and hippocampal volume alongside genetic and demographic information are also used as biomarkers; C) Subsampling stable MCI to overcome the class imbalance. Repeated random subsampling of the stable MCI group 100 times for Gaussian Processes based models; D) Partitioning of total variance explained into independent and shared variance explained for each biomarker; Gaussian Processes that allow just for main effects of biomarkers to be modelled are constructed by adding all the univariate squared exponential kernels corresponding to each biomarker. Full-order interactions between all biomarkers considered are captured through a single multivariate squared exponential kernel; Full set of statistics (sensitivity, specificity, accuracy, AUC) are computed for each replicate. Paired t-tests are conducted between accuracy scores stemming from main effects models and a model containing a multivariate kernel to assess if the increase in generalisation to unseen data is statistically significant.

Using the ADNI dataset, we analysed people with stable and progressive MCI to investigate: (i) the independent value each biomarker has in predicting conversion; and (ii) whether modelling nonlinear interactions between biomarkers could improve the fit of classification models that predict conversion to Alzheimer’s. We included multiple biological indices, to capture different facets of Alzheimer’s: APOE genotype, CSF measures of amyloid-β42, total tau and p-tau, PET measure of amyloid-β deposition, and structural MRI measures of global ‘brain age’ and of hippocampal volume.

## Material and methods

A high-level overview of the methods used in the study is included in Figure 1.

### Participants

Participants were drawn from the publicly-available ADNI dataset. Inclusion criteria included the availability of all biomarkers (genetic, fluid, PET, MRI), a diagnosis of MCI at baseline imaging assessment and the availability of clinical follow-up three years after the imaging assessment to determine disease progression. Stable MCI participants still met the criteria for MCI after three years; progressive MCI participants met the criteria for a clinical diagnosis of dementia at or before the three-year assessment. In total, n = 206 people with MCI were included in the analysis; stable MCI n = 158 (age range 55-89 years, median age=71.60 years), progressive MCI n = 48 (age range 55-84 years, median age = 73.85 years). Further participant details are included in Table 1.

**Table 1.**
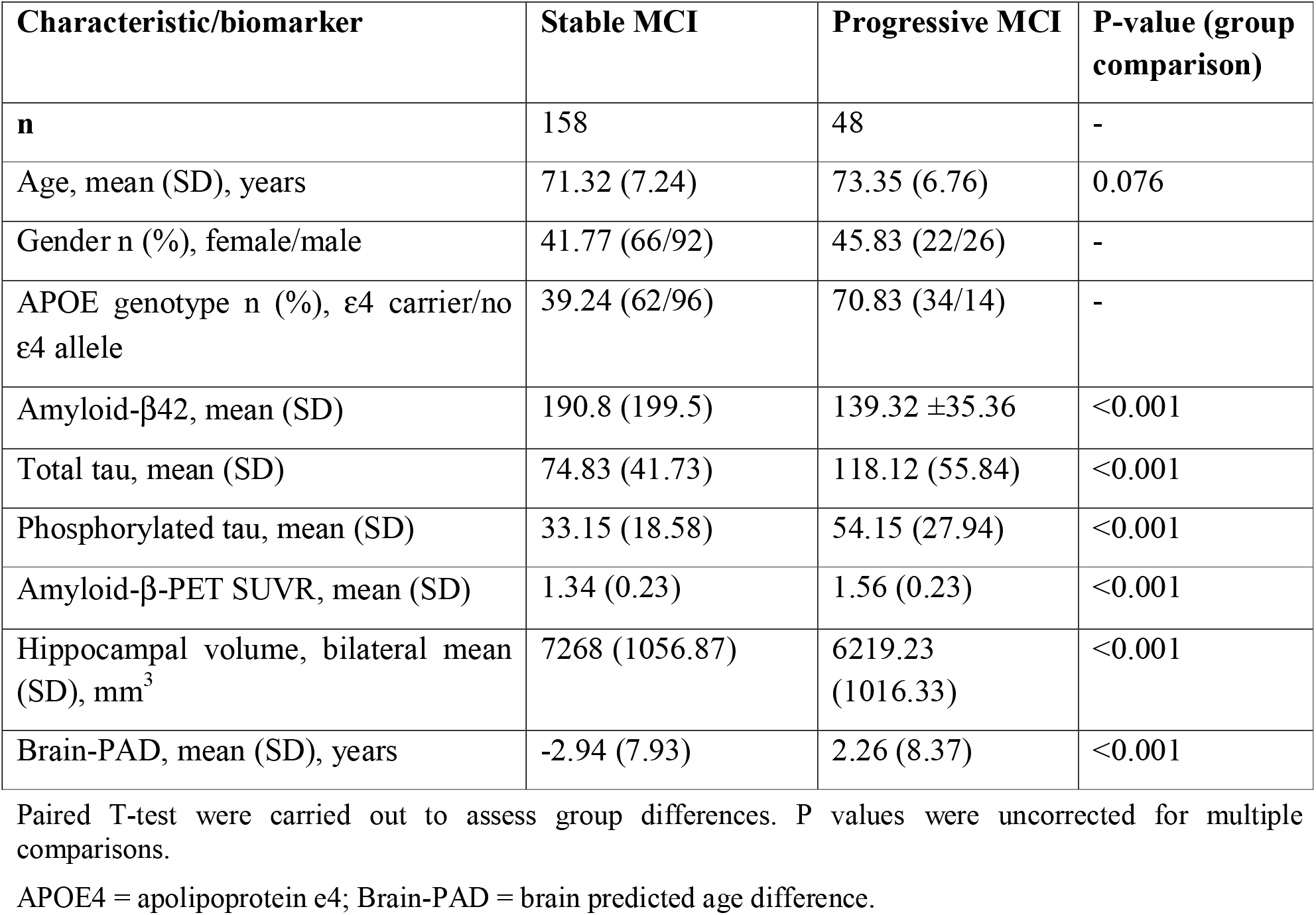
Participant characteristics and biomarker values

| Characteristic/biomarker | Stable MCI | Progressive MCI | P-value (group comparison) |
| --- | --- | --- | --- |
| **n** | 158 | 48 | - |
| Age, mean (SD), years | 71.32 (7.24) | 73.35 (6.76) | 0.076 |
| Gender n (%), female/male | 41.77 (66/92) | 45.83 (22/26) | - |
| APOE genotype n (%), ε4 carrier/no ε4 allele | 39.24 (62/96) | 70.83 (34/14) | - |
| Amyloid-β42, mean (SD) | 190.8 (199.5) | 139.32 ±35.36 | <0.001 |
| Total tau, mean (SD) | 74.83 (41.73) | 118.12 (55.84) | <0.001 |
| Phosphorylated tau, mean (SD) | 33.15 (18.58) | 54.15 (27.94) | <0.001 |
| Amyloid-β-PET SUVR, mean (SD) | 1.34 (0.23) | 1.56 (0.23) | <0.001 |
| Hippocampal volume, bilateral mean (SD), mm<sup>3</sup> | 7268 (1056.87) | 6219.23 (1016.33) | <0.001 |
| Brain-PAD, mean (SD), years | -2.94 (7.93) | 2.26 (8.37) | <0.001 |

Paired T-test were carried out to assess group differences. P values were uncorrected for multiple comparisons.
APOE4 = apolipoprotein e4; Brain-PAD = brain predicted age difference.

ADNI is a longitudinal observational study investigating whether serial brain imaging, clinical, and neuropsychological assessments can be combined to measure the progression of AD. Detailed information regarding the ADNI study’s procedures, including participant inclusion and exclusion criteria, and the complete study protocol can be found at http://adni.loni.usc.edu/. ADNI was launched in 2003 as a public-private partnership, led by Principal Investigator Michael W. Weiner, MD. ADNI is the result of efforts of co-investigators from a broad range of academic institutions and private corporations with participants being recruited from over 50 sites across the U.S and Canada.

To date, there are over 1500 adults, aged between 55 and 90, that participated in its three studies (ADNI-1, ADNI-GO, ADNI-2). The primary goal of ADNI has been to test whether serial MRI, PET and other biological markers, and clinical and neuropsychological assessment could be combined to measure the progression of MCI and early Alzheimer’s disease.

### Biomarker analysis

#### Biomarkers: Genetics

APOE genotype was determined from a 10ml blood sample taken during study screening and sent overnight to the University of Pennsylvania ADNI Biomarker Core laboratory for analysis. The APOE genotype of each participant was recorded as a pair of numbers indicating which two alleles were present; either ε2, ε3 or ε4.

#### Biomarkers: Fluid

Samples of 20ml of CSF were obtained from participants by a lumbar puncture with a 20- or 24-gauge spinal needle, around the time of their baseline scan. All samples were sent same-day on dry ice to the University of Pennsylvania ADNI Biomarker Core laboratory. There, levels of the proteins (amyloid-β42, total tau, and p-tau) were measured and recorded, as described previously(Shaw *et al*., 2009). By design, only a subset of ADNI participants had measurement of CSF levels.

#### Biomarkers: PET

Cerebral amyloid-β deposition was indexed using the PET tracer [18^F^]florbetapir (F-AV45). PET imaging was performed within two weeks of the baseline clinical assessments, as described previously (Jagust *et al*., 2015). In brief, four late-time five-minute frames are co-registered and averaged. The resulting image is converted to a 160×160×96 voxel static image with voxel dimension of 1.5mm^3^. Finally, an 8mm full-width half-maximum Gaussian kernel was applied (corresponding to the lowest resolution scanner used in the study). These primary data were downloaded from the ADNI database and used in the subsequent analyses.

F-AV45 data were nonlinearly registered into Montreal Neurological Institute 152 space (MNI152 space) using DARTEL (Ashburner, 2007). Initially the structural MRI images were segmented into grey matter and white matter using the Statistical Parametric Mapping (SPM12) software package (University College London, UK) and registered to a group average template. The group average template was then registered to MNI152 space. The F-AV45 standardized uptake value ratio (SUVr) image for each participant was registered to the corresponding T1-weighted MRI using a rigid-body registration. Finally, the individuals’ DARTEL flow field and template transformation was applied without modulation resulting in F-AV45 images in MNI152 space. The normalised maps were spatially smoothed (8mm full-width half-maximum Gaussian kernel). A neuroanatomical atlas (Tziortzi *et al*., 2011) and a grey-matter probability atlas in MNI152 space were employed to calculate regional SUVr values. SUVr values were quantified using the grey matter cerebellum as the reference region (defined as the intersection between the cerebellum region from the anatomical atlas and the grey-matter atlas, thresholded at p>0.5). The mean uptake value for cerebellar grey matter was obtained and each image was divided by this to generate an SUVr image for each participant. Finally, an average cortical SUVr value was obtained by calculating the mean SUVr value for all cortical regions (weighted by regional volume).

#### Biomarkers: Structural MRI

Three-dimensional T1-weighted MRI scans were acquired at either 1.5T (ADNI-1) or 3T (ADNI-2 and ADNI-GO) suing previously described standardized protocols at each site (Jack et al., 2008). All MRI scans were pre-processed using SPM12. This entailed tissue segmentation into grey matter and white matter, followed by a nonlinear registration procedure using DARTEL (Ashburner, 2007) to MNI152 space, subsequently followed by resampling to 1.5mm^3^ with a 4mm Gaussian smoothing kernel.

Processed grey matter and white matter images were then entered in the Pattern Recognition for Neuroimaging Toolbox (PRoNTo) software (Schrouff *et al*., 2013). Using a previously trained model that predicts chronological age from structural neuroimaging data, as per our previous work (Cole *et al*., 2015; Cole *et al*., 2017a; Cole *et al*., 2017c; Cole *et al*., 2018), we generated a brain-predicted age value for each participant. This step used a Gaussian Processes regression model with a linear kernel, with processed neuroimaging data as the independent variables, and age as the dependent variable. The training dataset used to define the model included n = 2001 health adults aged 18-90 years; further details have been reported previously (Cole *et al*., 2017b). Finally, we generated brain-PAD values; chronological age subtracted from brain-predicted age (i.e., the model predictions).

### Statistical analysis

#### Hierarchical partitioning of variance

We examined the independent contribution of each biomarker for distinguishing between stable and progressive MCI participants in a multivariate model, using hierarchical partitioning of variance (Chevan and Sutherland, 1991). Here, a hierarchical series of logistic regression models are fit, with all possible combinations of biomarkers. By comparing the variance explained by each model across all other models, an unbiased approximation of the variance attributable to each variable can be derived, both that independent of all other variables and that shared with other variables (due to the correlations between them).

Hierarchical partitioning of variance was carried out using the R package ‘hier.part’ with the following variables: age, sex; genetics (APOE4 genotype); CSF (amyloid-β42-CSF, total tau, p-tau); PET (amyloid-β-PET); structural MRI (hippocampal volume, brain-PAD). The number of variables was limited to nine as per Olea and colleagues (2010) to avoid inconsistent results. Thus, robust estimates of independent variance explained are obtained for each biomarker, which enabled assessment of the utility of including each biomarker when adjusting for all other biomarkers.

#### Multivariate biomarker models

We considered multiple statistical models, all with group (i.e., stable or progressive MCI) as the outcome (i.e., dependent) variable and with age, sex and APOE genotype as covariates. Each of the six imaging or fluid biomarkers were evaluated separately, alongside three different multivariate models, and a full model containing all biomarkers:

i. Hippocampal volume + brain-PAD + amyloid-β42-CSF
ii. Hippocampal volume + brain-PAD + amyloid-β-PET
iii. Hippocampal volume + amyloid-β-PET + p-tau
iv. Full model (hippocampal volume + brain-PAD + amyloid-β42-CSF + p-tau + total tau + amyloid-β-PET)

#### Gaussian Processes classification

Gaussian Processes are a supervised learning algorithm, that has found applications in many fields including our previous neuroimaging research (Cole *et al*., 2015), due to its Bayesian and non-parametric properties. The principal behind Gaussian Processes is that a measured (dependent) variable is modelled by defining a multivariate Gaussian distribution, hence we can view them as being a distribution over functions (Rasmussen and Williams, 2006).

To obtain Gaussian Process models where our predictor variables are independent, we assigned to each biomarker an individual squared exponential kernel, and subsequently all kernels are added together to derive a global kernel. To introduce interactions between biomarkers, we used all the covariates within the same squared exponential kernel. Duvenaud and colleagues (2011) proposed that this kernel construction defines an *m*-order interaction over covariate space, where *m* is the total number of covariates used in building the kernel. For example, two-way interactions were constructed by assigning the respective two biomarkers to a single kernel, subsequently adding this to the list of univariate kernels specific to each biomarker. For all models we used the non-sparse version of the Scalable Variational Gaussian Processes Classifier (Hensman *et al*., 2015).

#### Bootstrapping to adjust for class imbalance

The two groups of MCI patients were of differing size (progressive MCI n = 48, stable MCI n = 158), resulting in a class imbalance. To counter this imbalance, we subsampled the majority class (i.e., stable MCI) to generate a balanced dataset. To reduce the impact of sampling bias on this approach, we used bootstrapping (100 iterations) to randomly subsample the stable MCI group to create 100 different subsets, each compared with the entire progressive MCI group. Importantly, within each bootstrap, we used 10-fold stratified cross-validation to generate classification predictions for each participant. Sensitivity, specificity, (balanced) accuracy, area under the curve (AUC) and log likelihood values were calculated for each bootstrapped dataset. Values were then averaged across the 100 bootstrapped iterations.

#### Model comparisons

Two-way or three-way interactions were assessed by their relative improvement in log likelihood values compared to their corresponding main effects model. A paired t-test is used to test for differences, with the significance level set at 0.05. A list of the two-way and three-way interactions tested can be found in Table 2. To determine whether Gaussian Processes for classification resulted in improvements over less complex linear models, we performed the same classification task using logistic regression, with the same combination of different input variables and interaction terms (Supplementary Table S2). When considering model performance, we opted to focus on sensitivity to detecting progressive MCI as the chief criteria, under the assumption that a false-negative classification of stable MCI, is clinically more deleterious than a false positive.

**Table 2.** Biomarker model performance for predicting conversion from MCI to Alzheimer’s

| Model | Predictors | Sensitivity | Specificity | Balanced accuracy | AUC | T-test log likelihood, P-value |
| --- | --- | --- | --- | --- | --- | --- |
| **Individual biomarkers** |  |  |  |  |  |  |
| | Amyloid-β42-CSF | 0.625 | 0.895 | 0.760 | 0.749 | -51.10 |
|  | Total tau | 0.708 | 0.625 | 0.671 | 0.730 | -58.88 |
|  | P-tau | 0.729 | 0.666 | 0.697 | 0.722 | -58.57 |
| | Amyloid-β-PET | 0.666 | 0.666 | 0.666 | 0.729 | -58.09 |
|  | Hippocampal volume | 0.666 | 0.666 | 0.666 | 0.739 | -57.84 |
|  | Brain-PAD | 0.645 | 0.604 | 0.625 | 0.637 | -63.361 |
| **Main effects** |  |  |  |  |  |  |
| 1 | Hippocampal volume, brain-PAD, amyloid-β42 | 0.712 | 0.842 | 0.777 | 0.866 | -39.47 |
| 2 | Hippocampal volume, brain-PAD, amyloid-β-PET | 0.756 | 0.754 | 0.755 | 0.830 | -45.44 |
| 3 | Hippocampal volume, amyloid-β-PET, P-tau | 0.770 | 0.762 | 0.766 | 0.830 | -45.44 |
|  | Full model | 0.770 | 0.729 | 0.760 | 0.838 | -46.58 |
| **Two-way interactions** |  |  |  |  |  |  |
| 1 | Brain-PAD * amyloid-β42 | 0.708 | 0.848 | 0.778 | 0.864 | -39.45, 0.797 |
| | Amyloid-β42 * hippocampal volume | 0.713 | 0.846 | 0.780 | 0.865 | -39.08, <0.001 |
| 2 | Brain-PAD * amyloid-β-PET | 0.759 | 0.749 | 0.754 | 0.826 | -45.39, 0.46 |
| | Hippocampal volume * amyloid-β-PET | 0.723 | 0.788 | 0.756 | 0.830 | -44.60, <0.001 |
| 3 | Amyloid-β-PET * P-tau | 0.771 | 0.756 | 0.764 | 0.826 | -45.26, 0.015 |
| | Hippocampal volume * amyloid-β-PET | 0.733 | 0.806 | 0.769 | 0.840 | -43.02, <0.001 |
|  | P-tau * hippocampal volume | 0.758 | 0.768 | 0.763 | 0.826 | -45.114, 0.003 |
| **Three-way interactions** |  |  |  |  |  |  |
| 1 | Hippocampal volume * brain-PAD * amyloid-β42 | 0.724 | 0.830 | 0.777 | 0.861 | -39.422, 0.745 |
| 2 | Hippocampal volume * brain-PAD * amyloid-β-PET | 0.738 | 0.791 | 0.764 | 0.827 | -44.712, <0.001 |
| 3 | Hippocampal volume * amyloid-β-PET * P-tau | 0.748 | 0.812 | 0.780 | 0.835 | -43.232, <0.001 |

AUC = Area Under Receiver Operator Characteristic Curve. T-tests (paired) based on comparison between accuracy and log likelihood scores on 100 bootstrapped sets are provided as a mean to assess the relative increase in generalisation attributable to the introduction of either two-way interactions or three-way interactions between biomarkers in comparison to main effects only variants.

### Data availability

All data used for the preparation of this article was obtained from ADNI and are publicly-available. For up-to-date information, see www.adni-info.org. Our statistical analysis code is available online (https://github.com/SebastianPopescu/nonlinear_interaction_GPs).

## Results

A total of n = 158 stable MCI participants and n = 48 progressive MCI participants were included in our study. The two different groups are well matched in age and gender. In terms of APOE4 status, there is a significant difference, with 70.83% of progressive MCI being APOE4 positive, in comparison with 39.24% for stable MCI. Group differences were observed for all other biomarkers (Table 1).

### Biomarkers capture independent facets of Alzheimer’s progression risk

Hierarchical partitioning of variance was used to determine unique contributions of different predictor variables as well as their shared contribution towards prediction of conversion from MCI to Alzheimer’s after three years. The total variance explained by this additive model was R^2^ = 0.51, *P* < 0.001. Independent R^2^ values were: age = 0.028, sex = 0.004, APOE4 = 0.069, amyloid-β42-CSF = 0.063, total tau = 0.054, p-tau = 0.045, amyloid-β-PET = 0.048, hippocampal volume = 0.126, brain-PAD = 0.076. A substantial proportion of the variance was shared between variables, indicating that these biomarkers reflect some common as well as unique processes underlying progression from MCI to Alzheimer’s (Figure 2).

**Figure 2.**
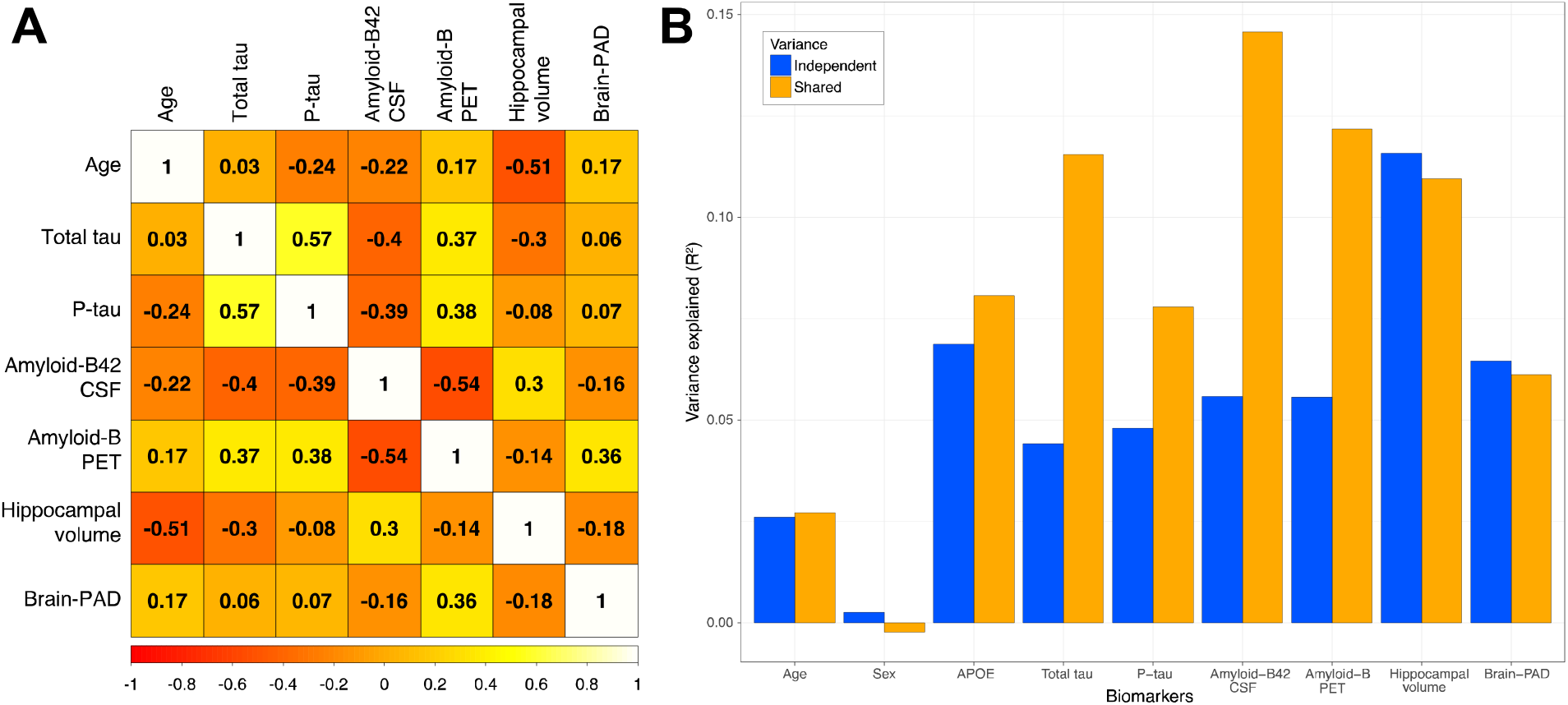
Independent and shared variance across biomarkers in predicting conversion to Alzheimer’s. A) Correlation matrix of numerical biomarkers, showing Pearson’s r values in balanced sub-sample of stable and progressive MCI participants (n = 96). B) Estimates of independent and shared variance for individual biomarkers in a logistic regression model with conversion to Alzheimer’s labels as the outcome variable and age, sex, APOE genotype, total tau, P-tau, amyloid-β42-CSF, amyloid-β-PET, hippocampal volume and brain-PAD as predictors. Estimates derived from hierarchical partitioning of the variance, with blue bars indicating independent (i.e., unique) variance attributable to each biomarker and yellow indicating variance shared with one or more other biomarkers.

### Logistic regression does not benefit from interaction terms

When using logistic regression to classify stable and progressive MCI patients, performance was moderately accurate, with balanced accuracies ranging from 0.67-0.72 (see Supplementary Table S2). Notably, for all models, classification sensitivity (range 0.61-0.65) was substantially lower than specificity (range 0.86-0.88), indicating that the logistic regression was better at identifying stable MCI patients than progressive MCI patients. When comparing the models containing interactions terms with their additive counterparts, t-tests showed than none of the interaction models were significant. This indicates that using standard logistic regression methods, where interactions are linear, there is no improvement in classification performance when including interaction terms.

### Nonlinear interactions between hippocampus and amyloid-based biomarkers

Different models were compared to assess the influence of including nonlinear interactions when classifying stable from progressive MCI (Table 2). For the models using biomarkers separately, P-tau showed the best sensitivity (0.729), while amyloid-β42-CSF had the highest accuracy (0.760). However, the AUC values for all these independent measures was broadly equivocal (AUCs range 0.722-0.749), except for brain-PAD, which was lower (AUC = 0.637). For the additive models of main effects, sensitivity ranged from 0.712-0.770, including the full model containing all biomarkers. When adding interaction terms for Model 1, we found that a bivariate interaction between hippocampal volume and amyloid-β42-CSF significantly improves model fit (i.e., significantly lowers log-likelihood). For Model 2, a two-way interaction between hippocampal volume and amyloid-β-PET also improved model fit. For Model 3, all three interactions improved model fit. That is, amyloid-β-PET by P-tau; amyloid-β-PET by hippocampal volume; P-tau by hippocampal volume. Finally, the inclusion of a three-way interaction between hippocampal volume, amyloid-β-PET and CSF P-tau also improved model fit for Model 2 and Model 3, but not Model 1. Notably, the sensitivity (range 0.708-771) and specificity (range 0.749-0.848) of the interaction models was similar to those of the additive main-effect models, despite the reductions in log-likelihood.

### Visualising nonlinear interactions: contour plots

The greater performance improvement occurred when including a two-way nonlinear interaction between amyloid-β-PET and hippocampal volume when adjusting for P-tau (Model 3). To investigate the relationships between these three biomarkers we used ‘contour’ plots to compare the decision boundaries between the main effects model, including interaction terms (Figure 3). A decision boundary can be defined as the point in measured space at which the classification is equivocal, in other words where there is greatest statistical uncertainty about which class (stable MCI or progressive MCI) an individual belongs to. The further away from the decision boundary, the more certain the model is that an individual is either stable MCI or progressive MCI, depending on the direction. In Figure 3, red regions are points in space with a high certainty of being progressive MCI, blue regions the equivalent for stable MCI. The decision boundaries are the white-coloured regions where the blue and red contours converge. These contour plots are analogous to topographical maps of mountainous terrain; strong colours represent mountain peaks, while white areas represent valleys, the decision boundaries.

**Figure 3.**
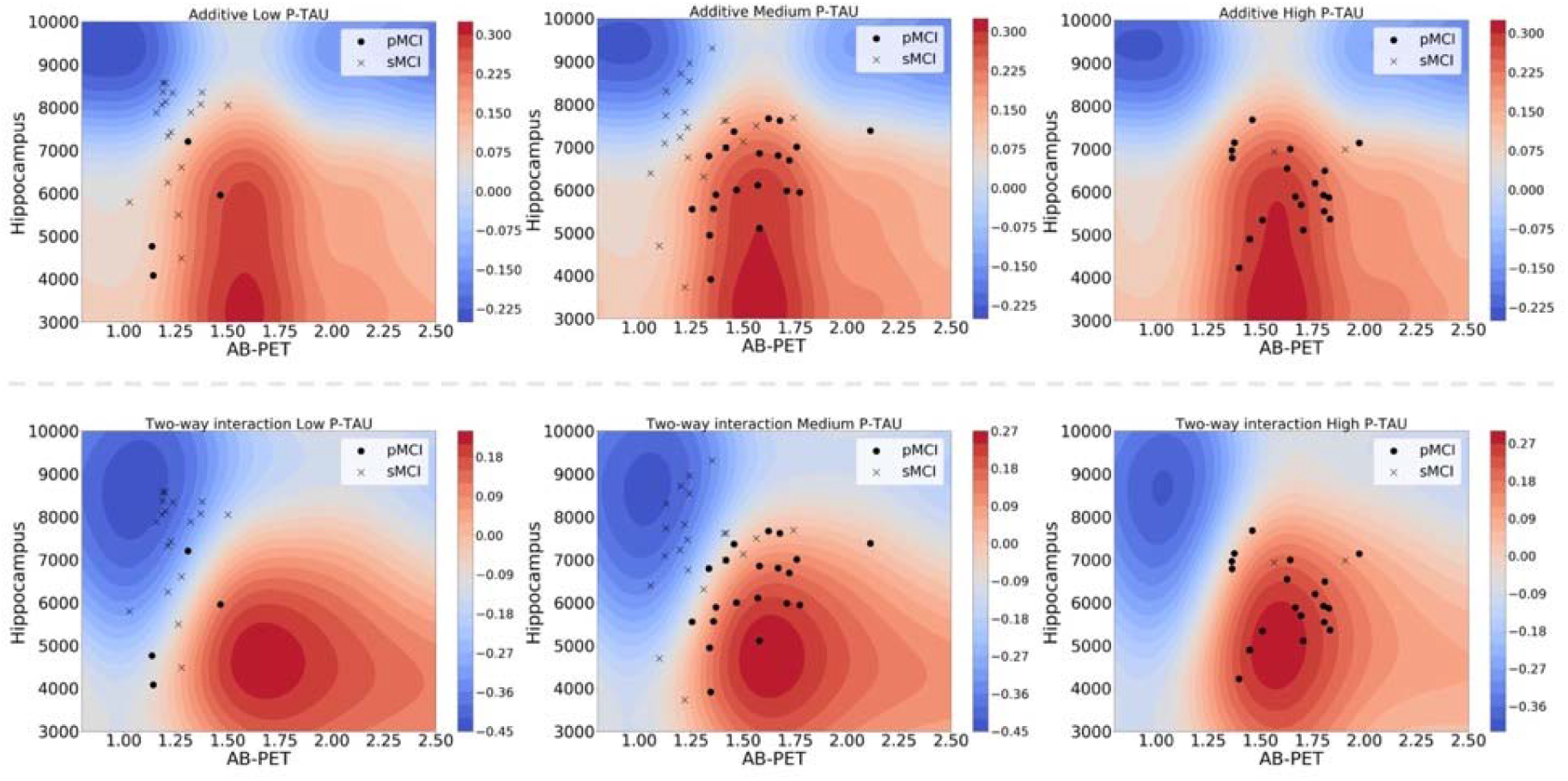
Decision boundaries for the hippocampal volume, amyloid-β-PET and P-tau classification model. Contour plots illustrate the decision boundaries for different classification models (Data based on Model 3). Positive values (red contours) indicate regions where the probability of being classified as progressive MCI is increased; negative values (blue contours) where that probability is decreased. Scatterplots of amyloid-β-PET (x-axis) and hippocampal volume (y-axis) are overlaid (crosses = stable MCI, dots = progressive MCI), with each column representing a tertiary split of participants based on P-tau values (<22.22 = low P-tau, 22.22-36.25 = medium P-tau; >36.25 = high P-tau). A) Additive model (no interactions). First row captures the dynamics of the summation of the univariate kernels associated to each biomarker, as P-tau is increased from. B) Two-way interaction model. Values on this row represent how much information towards prediction the interaction term is adding on top of the univariate kernels.

From inspection of the contour plots it is evident that for different levels of P-tau (based on this illustrative tertiary split), the relationship between hippocampal volume and amyloid-β-PET changes. In particular, for participants with medium P-tau levels, the contours are a better fit to the distribution of stable (represented by crosses on Figure 3) and progressive MCI participants (represented by dots) for the interaction model compared to the additive model. In practice, an individual patient could be located in this biomarker space, by using measurements of P-tau, hippocampal volume and amyloid-β-PET. A clinician could use this plot gain additional understanding of whether an individual patient more closely resembles a stable or progressive MCI patient, or whether they sit near the decision boundary. The ‘sharper’ the contrast between the red and blue contours, the less uncertainty there is in a classification decision, as is the case for the nonlinear compared to the additive model depicted in Figure 3.

## Discussion

Here, we modelled nonlinear interactions between a panel of biomarkers (neuroimaging, genetic, CSF) to predict disease progression from MCI to Alzheimer’s disease. Statistical models that included nonlinear interaction term explained conversion risk moderately better than additive linear models (in terms of model fit), though the influence of these interaction models of the sensitivity to progressive MCI was negligible. Importantly, our panel of biomarkers all independently explained a proportion of the variance in a logistic classification model. Though perhaps unsurprising, given evidence of the influence of these biomarkers in Alzheimer’s (Jack *et al*., 1999; Gaser *et al*., 2013; Xu *et al*., 2013; Caminiti *et al*., 2018), this justifies the inclusion of different sources of information, despite the evident shared variance (i.e., bivariate correlations) between biomarkers. This is even the case for measures of amyloid; whereby the PET-derived and CSF-derived measures were correlated r = -0.54. Crucially, our results show that a proportion of the non-shared variance between these two biomarkers still relates to disease progression. This is in line with previous findings (Mattsson *et al*., 2014), which showed that amyloid-β42-CSF and amyloid-β-PET provide partially independent information about a wide range of Alzheimer’s measures. While a practitioner’s decision about whether to acquire PET or CSF measures of amyloid for a single patient are likely influenced by cost and invasiveness, statistically including both will improve predictive accuracy.

We found that the nonlinear interaction between amyloid-β-PET and hippocampal volume significantly increases our model fit on unseen data, suggesting an element of synergy between the two biomarkers. By inspecting the model’s ‘contours’ (Figure 3), we observe that this increased model fit is attributable to ‘sharper’ decision boundaries. The probability of conversion for people with low hippocampal volume and high amyloid-β-PET values, was higher when compared to probability values obtained from a model where the biomarkers are treated independently, suggesting that modelling interactions explains more variance and decreases statistical uncertainty. Our finding concurs with the results from Bilgel and colleagues (2018), where an interaction was found between hippocampal volume and amyloid-β-PET in predicting cognitive decline, in their study of n = 171 older adults from the Baltimore Longitudinal Study of Aging. Additionally, we found evidence that a three-way interaction between amyloid-β-PET, hippocampal volume and P-tau further improves model fit compared to a main-effects model, suggesting a three-way interaction between these biomarkers. We also found evidence of nonlinear interactions between hippocampal volume and: CSF P-tau, amyloid-β-PET, amyloid-β42-CSF. This supports the idea that there is a ‘threshold effect’ on hippocampal volume, whereby larger hippocampal volumes are associated with stable MCI, irrespective of changes in other biomarkers, while with smaller hippocampal volumes, the readouts of other biomarkers are more predictive of progressive MCI. Potentially, the presence of intact hippocampal networks enables functional compensation to ameliorate the deleterious nature of amyloid and tau deposition, however, once hippocampal atrophy crosses a certain threshold, the consequences of abnormal protein deposition become clinically manifest.

Due to the nonparametric nature of Gaussian Processes, prediction probabilities in biomarker ‘space’ can be illustrated using contour plots, which provide visual information about the relationship between groups of biomarkers. When examining the contour plots and respective decision boundaries for Model 1, we observed high statistical ‘certainty’ in classifying participants as stable MCI in the case of high amyloid-β-CSF values, more specifically values above 192pg/mL. Interestingly, this corresponds to the threshold proposed by Shaw and colleagues (2009), who also used ADNI. Models including high amyloid-β-CSF achieved high levels of specificity, with the individual amyloid-β-CSF model having a specificity of 0.895. This means that amyloid-β-CSF is useful for indicating that an individual is stable MCI, but given the sensitivity of 0.625, is less valuable in identifying cases of progressive MCI.

The performance of our Gaussian Processes method for predicting MCI conversion was comparable to previous approaches (see Rathore *et al*., 2017 for review). For example, Cheng and colleagues (2015) used a multimodal manifold-regularized transfer learning with semi-supervised learning on MRI, PET scans and CSF features to achieve AUC = 0.852 (accuracy = 80.1%, sensitivity = 85.3%, specificity = 73.3%). Korolev and colleagues (2016) used the probabilistic multiple kernel learning classifier, obtaining AUC = 0.87 (accuracy = 80%, sensitivity = 83%, specificity = 76%). Moradi and colleagues (2015) achieved an AUC of 0.90 (accuracy = 82%, specificity = 74%, sensitivity = 87%) using a univariate structural MRI biomarker alongside a variety of cognitive scores. Besides these, Hor and colleagues (2016) implemented a Scandent tree approach on T1-weighted MRI ROI and global measures stemming from both FDG-PET and AV45-PET scans (accuracy = 0.815, sensitivity = 0.831, specificity = 0.803, AUC = 0.872). In the current analysis, our best performing model (in terms of sensitivity to progressive MCI) reached sensitivity = 0.771, specificity = 0.756, accuracy = 0.764, AUC = 0.826. Direct comparison of these values is complicated by the varying sample sizes in each study, generally based on biomarker availability (e.g., fewer participants had amyloid-PET scans). Our model performance results are sufficiently accurate to enable reasonable scrutiny of relationship between variables. Nevertheless, no published model to date has reach sufficiently high sensitivity to warrant clinic adoption for the prediction of developing Alzheimer’s in people with MCI, where values of 95% or greater are likely to be necessary to influence clinical decision making.

Our study has some important strengths and weaknesses. Ours is the first study to model nonlinear interactions between biomarkers in predicting MCI conversion to Alzheimer’s, properly utilising the available information in a more biologically-valid manor. Here, we focused on model prediction, unlike previous work on interactions in MCI conversion (Fortea *et al*., 2014; Pascoal *et al*., 2017b), that relied on making inferences based solely on p-values. Recently, Bzdok and colleagues (2018a) highlighted the diverging end results between classical statistics and machine learning, with the former drawing population level inferences from samples, whereas the latter is seeking to find generalisable predictive patterns. In further work, Bzdok and colleagues (2018b) showed that low p-values do not necessarily translate to generalisable biomarkers in out-of-sample data. Our model offers a robust and interpretable framework by which we can visualize how biomarkers dynamically interact and how those interactions affect the decision boundaries of classifying participants as stable MCI or progressive MCI. For example, this framework could be used in designing clinical trials to assess how much change in a given biomarker must occur for an individual to move to from being at low risk from conversion from MCI to Alzheimer’s to being at high risk (i.e., moving from blue regions to red in Figure 3).

Weaknesses of the current study include the limited sample size and lack of a replication dataset. While we did use ‘held-out’ data to test model generalisability, we were unable to test the predictive performance of our models to truly independent data. This is one key limitation of multi-modality prediction models, as beyond ADNI, datasets containing MRI, amyloid-PET and CSF measures are challenging to acquire and generally not openly-available to the research community. Another limitation is that we focused on the MCI stage of the disease. Evidence suggests that Alzheimer’s pathogenesis commences years prior to any cognitive symptoms, so future work should focus on younger, at-risk groups, in order to maximise the window for interventions prior to Alzheimer’s disease manifestation. Furthermore, the diagnostic stages defined in ADNI are made clinically, while the gold standard for AD diagnosis is post-mortem. Potentially, some of the clinical assessments are inaccurate.

In conclusion, our findings suggest that multiple underlying neurobiological processes both act independently and interact in a nonlinear fashion during progression from MCI to Alzheimer’s. By capturing atrophy (hippocampal volume), amyloid-β deposition (using PET) and neurofibrillary tangle formation (CSF P-tau) in a nonlinear interactive model, can better fit models to predict conversion than independent effects alone, though main-effects multi-modality model still offer reasonable performance. The impact of modelling nonlinear interactions implies that threshold effects and pathological ‘plateaus’ are present during disease progression and that measuring multiple biomarkers will be necessary to best predict outcomes. The current data do not suggest clinically meaningful benefits of nonlinear-interaction models yet, though our novel visualisation method (contour plots) could be helpful in clinical contexts and provides strong face validity for our approach. Finally, multi-faceted therapeutic interventions are likely to be necessary, as simply influencing the levels of a single biomarker will be insufficient to modify the disease in all individuals.

## Data Availability

https://github.com/SebastianPopescu/nonlinear_interaction_GPs

## Acknowledgements

Data collection and sharing for this project was funded by the Alzheimer’s Disease Neuroimaging Initiative (ADNI) (National Institutes of Health Grant U01 AG024904) and DOD ADNI (Department of Defense award number W81XWH-12-2-0012). ADNI is funded by the National Institute on Aging, the National Institute of Biomedical Imaging and Bioengineering, and through generous contributions from the following: AbbVie, Alzheimer’s Association; Alzheimer’s Drug Discovery Foundation; Araclon Biotech; BioClinica, Inc.; Biogen; Bristol-Myers Squibb Company; CereSpir, Inc.; Cogstate; Eisai Inc.; Elan Pharmaceuticals, Inc.; Eli Lilly and Company; EuroImmun; F. Hoffmann-La Roche Ltd and its affiliated company Genentech, Inc.; Fujirebio; GE Healthcare; IXICO Ltd.; Janssen Alzheimer Immunotherapy Research & Development, LLC.; Johnson & Johnson Pharmaceutical Research & Development LLC.; Lumosity; Lundbeck; Merck & Co., Inc.; Meso Scale Diagnostics, LLC.; NeuroRx Research; Neurotrack Technologies; Novartis Pharmaceuticals Corporation; Pfizer Inc.; Piramal Imaging; Servier; Takeda Pharmaceutical Company; and Transition Therapeutics. The Canadian Institutes of Health Research is providing funds to support ADNI clinical sites in Canada. Private sector contributions are facilitated by the Foundation for the National Institutes of Health (www.fnih.org). The grantee organization is the Northern California Institute for Research and Education, and the study is coordinated by the Alzheimer’s Therapeutic Research Institute at the University of Southern California. ADNI data are disseminated by the Laboratory for Neuro Imaging at the University of Southern California.

## Abbreviations

AUC: Area under the receiver operating characteristic curve
ADNI: Alzheimer’s Disease Neuroimaging Initiative
Brain-PAD: Brain-predicted age difference
MCI: Mild Cognitive Impairment
P-tau: phosphorylated-tau
SUVR: Standardized uptake value ratios

